# Oxytocin and its role in energy balance and metabolic factors: A living systematic review and meta-analysis protocol

**DOI:** 10.1101/2025.04.04.25325238

**Authors:** Alina I. Sartorius, Heemin Kang, Elisabeth Deilhaug, Kjersti Mæhlum Walle, Elizabeth A. Lawson, Kamryn Eddy, Dennis van der Meer, Lars T. Westlye, Daniel S. Quintana

## Abstract

Oxytocin is a neuropeptide that first received research attention for its role in parturition and lactation and has since been implicated in various cognitive and behavioral processes. More recently, evidence has shown that oxytocin is also involved in additional physiological domains, such as energy balance and metabolism. To our knowledge, no recent meta-analysis on the effects of exogenous oxytocin on energy balance and metabolism has been conducted. With this article, we present a protocol that describes the plan for a living systematic review and meta-analysis investigating the role of oxytocin in energy regulation and metabolism, as well as the variables which may moderate this potential effect.

## Background

Oxytocin is a neurotransmitter and hormone predominantly synthesized in the paraventricular and supraoptic nuclei of the hypothalamus (Ivell, 1986) that was first known for its physiological functions in the contraction of the uterus during parturition and the milk letdown reflex in offspring feeding (Howarth & Botha, 2001; Ruis et al., 1981). Only during the late 1990s and early 2000s has its role in cognitive and social domains been recognized (Lippert et al., 2003). Apart from the discovery of its cognitive and social properties, recently, studies have been accumulating to support the notion that oxytocin is also involved more broadly in energy regulation and metabolism (Jurek & Neumann, 2018; Lawson, 2017; McCormack et al., 2023; Winterton et al., 2020).

There are various ways that oxytocin’s effects on energy regulation and metabolism have been operationalised, which can be broadly categorised as feeding behavior, body weight and composition, and biological markers of metabolic regulation, such as glucose homeostasis.

In the domain of feeding behavior, a single intranasal administration of oxytocin led to a reduction in food cravings in healthy, typical-weight adult women (Striepens et al., 2016). Further, intranasal oxytocin has been shown to reduce homeostatic and reward-driven food intake in humans (Burmester et al., 2018; Lawson et al., 2015; Ott et al., 2013; Plessow et al., 2024; Thienel et al., 2016).

The anorexigenic effects of oxytocin on body weight and body composition have been studied thoroughly in the past decades in animal models (e.g., (Blevins et al., 2015; Morton et al., 2012). In humans, in 2021, Espinoza and colleagues investigated the role of oxytocin administered via the intranasal route in sarcopenic obesity, showing an increase of lean body mass in elderly adults (Espinoza et al., 2021). On the other hand, a study on the effects of intranasal oxytocin on weight loss did not report statistically significant evidence for an anorexigenic effect of oxytocin in individuals with hypothalamic obesity, at least for a 16-24 International Unit dose (McCormack et al., 2023). While a pilot study demonstrated weight loss with intranasal oxytocin in adults with obesity (Zhang et al., 2013), a larger clinical trial did not show effects on weight or body composition (Plessow et al., 2024).

In terms of biological markers, studies in animal models have mostly demonstrated beneficial effects (e.g., improved glucose tolerance (Deblon et al., 2011; Zhang et al., 2013)), whereas effects in humans have been more inconsistent (Lawson et al., 2020). For instance, research has reported that in human males, a single dose of intranasal oxytocin (compared to a placebo) has been linked to insulin suppression and reduced glucose concentrations after meal-intake in normal weight but not obese men (Brede et al., 2019).

In 2021, one team of researchers conducted a systematic review and meta-analysis investigating the effects of intranasal oxytocin on food intake and cravings in overweight individuals, those without psychiatric conditions, and those with an eating disorder (Chen et al., 2021). However, this review did not account for publication bias nor effect size dependency, which can lead to inaccurate summary effect size estimates. Leslie and colleagues (2018) similarly reported a meta-analysis on oxytocin and energy intake, which only accounted for small study bias, rather than publication bias (Leslie et al., 2018). To date, there have been numerous other published reviews describing the literature supporting the important role of oxytocin in eating behavior and metabolism, but none have included formal data synthesis. Moreover, findings on metabolic effects of oxytocin have been inconclusive.

To address this gap in synthesized data in the literature, we plan to perform a systematic review and meta-analysis to better understand the effects of exogenous oxytocin on energy balance and different metabolic factors, respectively, using state-of-the-art correction methods to detect and account for publication bias. We will conduct three subgroup meta-analyses to capture the full complexity of aspects that constitute energy balance and metabolism. The meta-analyses will be grouped into subgroups based on a preliminary scoping review of the literature: a) food intake, b) measures of body weight and composition (e.g., obesity, lean muscle mass), and c) biological markers of energy regulation and metabolism (e.g., insulin sensitivity, serum glucose levels). To understand how different covariates, such as age, study population, or drug regimen might influence the variation in study results, we will also perform moderator analyses. A summarized form of this protocol will be made available on PROSPERO following its registration template.

We will investigate effects of exogenously administered oxytocin-based therapeutics versus a placebo in humans on:

1. Caloric intake
2. Body weight and composition
3. Biological markers of energy regulation and metabolism

**Table 1:** Population, intervention, comparator, outcome (PICO)

| Population | Intervention | Comparator | Outcome |
| --- | --- | --- | --- |
| Healthy and clinical human individuals | Oxytocin administration | Placebo | Behavioral and physiological measures of energy regulation and metabolism |

### Search procedure

We will follow the “Statement for Reporting Literature Searches in Systematic Reviews” from the Preferred Reporting Items for Systematic Reviews and Meta-Analyses – Living Systematic Reviews (PRISMA-LSR) guidelines when reporting the outcomes (Akl et al., 2024). The following databases will be used: PubMed, Web of Science, Embase, Cochrane Library, Scopus, and PsycInfo. Search terms were determined with help of an academic librarian from the University of Oslo library and customized to suit each database.

### Search terms

The following search terms will be utilized to implement the primary search. The three categories operationalise the three hypotheses and will each be combined with oxytocin/oxytocin* in three separate queries in the different search engines.

((oxytocin) OR (oxytocin*)) AND

1)

((food) OR (foods) OR (food intake) OR (nutritio*) OR (energy intake) OR (volume intake) OR (meal intake) OR (nutritional intake) OR (feeding, refeeding) OR (re-feeding) OR (eating) OR (diet*) OR (snack*) OR (caloric intake) OR (calorie intake) OR (caloric intake) OR (calories) OR (meal*) OR (appetit*) OR (satiety) OR (satiation) OR (fullness) OR (food craving) OR (feeding behavio?r) OR (food cue) OR (craving) OR (hunger) OR (starvation) OR (starving) OR (overeating) OR (underfeeding) OR (fasting) OR (hyperphagia) OR (polyphagia) OR (gustation) OR (olfaction) OR (taste) OR (smell) OR (aroma) OR (flavo?r))

2)

((obesity) OR (obese) OR (adipos*) OR (anorexi*) OR (anorectic) OR (bulimi*) OR (weight) OR (body weight) OR (bodyweight) OR (body composition) OR (body mass index) OR (BMI) OR (waist-to-hip ratio) OR (lean muscle mass) OR (lean body mass) OR (fat) OR (overweight) OR (underweight))

3)

((metaboli*) OR (energy expenditure) OR (REE) OR (energy spending) OR (spending energy) OR (using energy) OR (energy regulation) OR (energy intake) OR (energy sensing) OR (energy balance) OR (energy homeostasis) OR (energy needs) OR (leptin) OR (ghrelin) OR (insulin*) OR (HOMA-IR) OR (adiponectin) OR (lipolysis) OR (fat oxidation) OR (triglyceride*) OR (cholesterol) OR (glycerol) OR (glucose) OR (glucagon) OR (blood sugar) OR (OGTT) OR (lipogenesis) OR (lipoproteins) OR (free fatty acids) OR (ketones) OR (ketone bodies) OR (gastric emptying) OR (gastroparesis) OR (dyspepsia) OR (gastrointestin*) OR (diabet*) OR (hypoglycaemia) OR (hypoglycemia) OR (glucose deficiency))

### Inclusion and exclusion criteria

Studies will be *included* if they meet the following criteria:

1. Population: Both healthy and individuals from clinical populations will be considered
2. Intervention: Exogenous oxytocin administration via the nasal, intravenous, or oral route
3. Comparator: The effect of oxytocin was studied using within- or between-subject randomized placebo-controlled trials.
4. Outcome: Energy balance and metabolism operationalised through caloric intake, body weight/composition, and biological markers
5. Language: Publication in English

No restrictions regarding publication year will be applied.

Studies will be *excluded* if they meet any one of the following criteria:

1. Animal studies
2. A study investigating endogenous oxytocin
3. Case reports, cross-sectional studies, case-control studies, or cohort studies

## Data extraction and management

### Study selection

The titles and abstracts of the preliminary selection of studies detected through primary database searches will be screened for initial eligibility by the first author (AIS) based on the inclusion and exclusion criteria using Covidence (www.covidence.org). Subsequently, the full article of studies deemed eligible will be evaluated. Using a custom R script, a second author (ED) will assess a randomly selected 20% sample of these studies. Any discrepancies will be mediated by senior author DSQ. Following this, in a secondary search, the reference lists of studies from the first screening and articles citing studies from the first screening will be searched to identify more potential eligible studies. These studies will be screened following the same procedure as from the primary search. Lastly, all duplicates will be removed from the final list of eligible studies.

### Data extraction

First author (AIS) will extract data from each study to calculate standardized mean differences (this may include means, standard deviations, and *p*-values). Another author (HK) will extract data from a 25% sample of the eligible studies chosen randomly using a custom R script. Any discrepancies will be mediated by author (DSQ).

Furthermore, the following data will be extracted in addition:

- Publication year of the study
- Sample characteristics, including sample size, mean age, age range, healthy/clinical cohort and sex distribution
- Study design, including task or experiment information and outcome measure(s)
- Oxytocin administration features, including administration route, dose and frequency
- Peer-review status (i.e., peer-reviewed vs. preprint or thesis)

### Missing data

Wherever a study has cases of missing data required for inclusion in the meta-analysis, data will be requested directly from the study authors. These requests will be for either raw data or computed effect sizes. Alternative methods will be used to extract data (e.g., WebPlotDigitizer (Rohatgi, 2011), and calculate effect size from summary statistics (Borenstein et al., 2009)) wherever primary data could not be obtained.

### Meta-analysis updates

This meta-analysis search will be updated at least two years after the completion of the initial search. We commit to at least two updates, that will take place in approximately 2027 and 2029.

## Data quality and bias assessment

### Power analysis

Statistical power for all included studies for a range of hypothetical effect sizes of interest will be calculated with the `metameta` R package (Quintana, 2023). `metameta` is a tool to calculate study-level power for meta-analyses for a range of hypothetical possible effect sizes, given that the true effect size of a population is typically unknown.

### Risk of bias

Risk of bias will be estimated using “RoB 2: A revised tool for assessing risk of bias in randomized trials” (Sterne et al., 2019).

### Small study bias

Egger’s regression test (Mathur & VanderWeele, 2020) will be used to assess small study bias and contour-enhanced funnel plots will be used for visualization (Mathur & VanderWeele, 2020).

### Publication bias

The potential presence and impact of publication bias will be assessed by conducting a sensitivity analysis utilizing the R package `PublicationBias` (Mathur & VanderWeele, 2020). A Robust Bayesian meta-analysis will be performed to complement this, evaluating the relative degree of evidence for or against publication bias (Maier et al., 2023).

### Protocol deviations

Any protocol deviations will be reported in the manuscript and/or a supplementary document.

## Statistical analyses

As the default, conventional two-level random-effects meta-analysis will be calculated to account for within- and between-study variability. In case that multiple effect sizes are extracted from the same study, we will perform a three-level random-effects meta-analysis to account for between-effect-size variability within studies. If not stated otherwise, all statistical analyses will be conducted and results visualized with the statistical software R (R Core Team, 2021) in RStudio (Posit team, 2023). The R package `metafor` (Viechtbauer, 2010) will be used for the frequentist meta-analysis, `RoBMA` (Maier et al., 2023) for the robust Bayesian meta-analysis, `metameta` (Quintana, 2023) for the study-level power analysis, and `TOSTER` (Lakens, 2017) for equivalence testing. Bayes factors <1 will be interpreted as evidence favouring the null hypothesis, 1-3 will be interpreted as anecdotal evidence favouring the alternative hypothesis, >3 will be considered moderate evidence, and >10 as strong evidence for the alternative hypothesis (Quintana & Williams, 2018). As this is a living meta-analysis with repeated meta-analysis, a Bonferroni adjusted *p*-value of 0.017 will be used for the calculation of summary effects size to adjust for three planned meta-analyses.

### Summary effect size

In order to address potential variation in experiment designs, we will deploy distinct methods for within-subject and for between-subject studies to calculate the effect size for data synthesis, given the dependent nature of between-group mean differences. This approach will help mitigate effect size dependency, a potential issue when multiple outcome measures are reported for the same subject within a single study. For between-subject studies, we will summarise the standardized between-group mean difference (SMD), as Hedge’s *g*. If this effect estimate is not available, other effect estimates will be converted to Hedge’s *g*.

### Equivalence testing

Calculated effect sizes may not reach statistical significance despite the presence of a true underlying effect, which may be due to underpowered analyses. In order to explore this possibility (i.e., if the statistical non-significance of effect sizes is due to the absence of a true effect or due to a lack of power to detect it), equivalence tests will be performed. Equivalence testing allows for the rejection of effects that are larger than a smallest effect size of interest (SESOI; (Lakens, 2017). To inform the equivalence bounds, we will use the smallest effect size as determined by Glaser et al. (2023) which is based on different meta-analyses of oxytocin intervention studies corrected for publication bias, thus Hedge’s *g* = ±0.06 (Glaser et al., 2023). Given these rather conservative bounds and which effect sizes are otherwise typical to observe in the oxytocin research field (Kang et al., 2025; Quintana, 2020), we also consider the reported medium effect size for additional, less stringent bounds, Hedge’s *g* = ±0.023 (Glaser et al., 2023).

### Study heterogeneity

To account for and quantify the degree of between-study heterogeneity regarding outcome measures (e.g., effect sizes), Restricted Maximum Likelihood (REML; Hardy & Thompson, 1996) will be used as heterogeneity estimator to estimate τ2.

### Sample heterogeneity

A certain degree of heterogeneity between the included studies is to be expected. Between-study heterogeneity due to differences in effect size reporting from one study to another, or due to random error within studies, will be tested by the use of Cochran’s Q, I^2^, and τ2.

### Moderator analyses

A fraction of between-study heterogeneity may also be explained by covariates or moderator variables, respectively. Analysis for subgroups of studies is a tool to test the degree to which such covariates play a role in the unexplained study heterogeneity. Covariates will be defined *a priori*. The following covariates will be included:

1. Study design: Whether a study has a within-subject or between-subject study design may impact the effect that oxytocin may have on the outcome measure.
2. Study population: Whether a cohort had a psychiatric/neurodevelopmental/metabolic condition or is healthy/typical may impact the effect that administered oxytocin has on an individual (Macdonald & Feifel, 2013).
3. Sex: Oxytocin operates in a sexually dimorphic manner. Therefore, sex will be included as a covariate that may moderate the effect the independent variable may have on the dependent variable (Quintana et al., 2024).
4. Age: Although we expect mostly adult populations, the populations can consist of younger or older adults and in certain conditions children, which could influence the outcome.
5. Administration route: All types of administration routes will be considered in this meta-analysis. Yet, different routes may lead to different outcomes due to for instance differences in absorption time and rate, or metabolization (Quintana et al., 2021).
6. Dosage: Previous research has shown that dosage may impact the outcome measure (Quintana et al., 2017), thus, this will be considered as a covariate.
7. Administration frequency: Whether oxytocin is administered in a single or with multiple doses may influence the outcome.
8. Endpoint: Are the results collected objectively (e.g., blood sample markers) or subjectively (e.g., self-reported)?
9. Endpoint status: Was the endpoint a primary outcome measure or another type of outcome measure?
10. Peer review status: This meta-analysis will also include preprints and theses. The review status (peer-reviewed vs. non-peer-reviewed) may moderate the outcome.

## Discussion

This planned systematic review and meta-analysis will systematically investigate the effects of exogenously administered oxytocin on energy balance and metabolic factors, taking potential moderating variables into consideration. Individual studies within this domain have been published, however, with mixed results. A combined systematic review and meta-analysis of this scope, controlling for different types of biases such as publication bias, to the author’s knowledge, has not been conducted yet. In synthesizing existing evidence on this topic, the aim of this planned analysis is to provide a better overview and understanding of oxytocin’s role in different facets of energy regulation and metabolism in humans.

## Data Availability

Not relavant as this article describes a protocol.

## Data and code availability

Data will be made available for download on a dedicated Open Science Framework (OSF) repository.

R scripts used in the analyses with information on specific parameter settings and additional notes will be made available at the same OSF repository.

## Author contributions

AIS and DSQ conceived the meta-analysis study idea. AIS and DSQ contributed to the design of the systematic review and the data analysis plan. AIS wrote the first and revised drafts of the protocol, with DSQ, HK, ED, KMW, EAL, KE, DVDM, LTW and DSQ contributing to the first draft of the protocol. All authors approved the final draft of the protocol. DSQ takes responsibility for the contents of the protocol.

